# Tocilizumab for treatment of mechanically ventilated patients with COVID-19

**DOI:** 10.1101/2020.05.29.20117358

**Authors:** Emily C Somers, Gregory A Eschenauer, Jonathan P Troost, Jonathan L Golob, Tejal N Gandhi, Lu Wang, Nina Zhou, Lindsay A Petty, Ji Hoon Baang, Nicholas O Dillman, David Frame, Kevin S Gregg, Dan R Kaul, Jerod Nagel, Twisha S Patel, Shiwei Zhou, Adam S Lauring, David A Hanauer, Emily Martin, Pratima Sharma, Christopher M Fung, Jason M Pogue

## Abstract

**Background:** Severe COVID-19 can manifest in rapid decompensation and respiratory failure with elevated inflammatory markers. This presentation is consistent with cytokine release syndrome in chimeric antigen receptor T cell therapy, for which IL-6 blockade is approved treatment.

**Methods:** We assessed effectiveness and safety of IL-6 blockade with tocilizumab in a single-center cohort of patients with COVID-19 requiring mechanical ventilation. The primary endpoint was survival probability post-intubation; secondary analyses included an ordinal illness severity scale integrating superinfections. Outcomes in patients who received tocilizumab compared to tocilizumab-untreated controls were evaluated using multivariable Cox regression with propensity score inverse probability weighting (IPTW).

**Findings:** 154 patients were included, of whom 78 received tocilizumab and 76 did not. Median follow-up was 47 days (range 28-67). Baseline characteristics were similar between groups, although tocilizumab-treated patients were younger (mean 55 vs. 60 years), less likely to have chronic pulmonary disease (10% vs. 28%), and had lower D-dimer values at time of intubation (median 2.4 vs. 6.5 mg/dL). In IPTW-adjusted models, tocilizumab was associated with a 45% reduction in hazard of death [hazard ratio 0.55 (95% CI 0.33, 0.90)] and improved status on the ordinal outcome scale [odds ratio per 1-level increase: 0.59 (0.36, 0.95)]. Though tocilizumab was associated with an increased proportion of patients with superinfections (54% vs. 26%; p<0.001), there was no difference in 28-day case fatality rate among tocilizumab-treated patients with versus without superinfection [22% vs. 15%; p=0.42].

**Interpretation:** In this cohort of mechanically ventilated COVID-19 patients, tocilizumab was associated with a decreased likelihood of death despite higher superinfection occurrence. Randomized controlled trials are urgently needed to confirm these findings.

**KEY POINTS:** *Question:* Can therapy with the IL-6 receptor antagonist tocilizumab improve outcomes in patients with severe COVID-19 illness requiring mechanical ventilation?

*Findings:* In this observational, controlled study of 154 patients, receipt of tocilizumab was associated with a 45% reduction in the hazard of death, despite twice the frequency of superinfection (54% vs 26%), both of which were statistically significant findings.

*Meaning:* Tocilizumab therapy may improve survival in patients with COVID-19 illness requiring mechanical ventilation. These results can inform clinical practice pending the results of randomized clinical trials.

## INTRODUCTION

SARS CoV-2, the virus responsible for COVID-19, has caused a global pandemic with over 5 million infections and 300,000 deaths as of May 20, 2020. Up to 20% of patients with COVID-19 develop severe illness characterized by worsening dyspnea and the need for supplemental oxygen.^1^ Patients may further progress to respiratory failure, acute respiratory distress syndrome (ARDS), multi-organ dysfunction, and death. Hyperinflammation may contribute to this deterioration, resulting in elevations in C-reactive protein, ferritin, lactate dehydrogenase (LDH), D-dimer, and various pro-inflammatory cytokines including interleukin-6 (IL-6).^1–6^ This profile resembles that seen in cytokine release syndrome (CRS) associated with chimeric antigen receptor (CAR) T-cell therapy and hemophagocytic lymphohistocytosis.^2,3,7^ In CRS, IL-6 blockade with tocilizumab has resulted in rapid improvement in respiratory and hemodynamic parameters,^8^ and the United States Food and Drug Administration has approved its use for CAR T-cell associated severe or life-threatening CRS.

As a result, adjunctive therapy with either IL-6 receptor antagonists (tocilizumab, sarilumab), or IL-6 antagonists (siltuximab) has been proposed as treatment for severe, progressive COVID-19. While multiple case series have suggested a potential role for tocilizumab^9–11^ or siltuximab (preprint),^12^ these reports are hampered by incomplete reporting, short durations of follow-up, and lack of control groups. Furthermore, infection is a concern with IL-6 blockade and cases of viral myocarditis^13^ and candidemia^14^ with tocilizumab have been reported. As secondary infection has been associated with increased mortality in COVID-19,^4^ controlled data are necessary to evaluate the risks and benefits of these therapies.

At our institution, IL-6 blockade with tocilizumab is considered for patients with severe COVID-19 and suspected hyperinflammation based on rapidly worsening respiratory status and elevated inflammatory markers, with the majority of usage occurring in patients requiring mechanical ventilation. Using our COVID-19 Rapid Response Registry infrastructure, we performed an observational study of outcomes in patients with COVID-19 requiring mechanical ventilation, comparing those treated with tocilizumab with those who were not.

## METHODS

Within the Michigan Institute for Clinical and Health Research, we developed a COVID-19 Rapid Response Registry for clinical characterization of persons with SARS-CoV-2 infection. The Registry includes core items from the International Severe Acute Respiratory and Emerging Infection Consortium (ISARIC) Clinical Characterization Protocol.^15,16^ This analysis follows STROBE recommendations.^17^ Ethics approval was obtained by the Institutional Review Board of the University of Michigan.

### Study Population

Patients were eligible for inclusion in this analysis if they were admitted to Michigan Medicine from March 9-April 20, 2020 for severe COVID-19 pneumonia, had a reverse-transcriptase polymerase chain reaction positive SARS-CoV-2 test, and required invasive mechanical ventilation. Follow-up continued through May 19, 2020. Patients were excluded if they were younger than 16 years, were intubated for conditions unrelated to COVID-19, or were enrolled into a randomized controlled trial (RCT) for sarilumab. This analysis focuses on comparative outcomes of mechanically ventilated patients who received tocilizumab and those who did not. Untreated patients who died prior to the opportunity to receive tocilizumab treatment per institutional criteria (within 48 hours of intubation) were excluded to minimize immortal time bias.^18^

### Tocilizumab exposure

Criteria for tocilizumab usage were developed by the institutional Antimicrobial Stewardship Program and Division of Infectious Diseases. Guidance was slightly modified during the study period based on drug availability, whether an IL-6 inhibitor clinical trial (sarilumab) was active, and experiences of the treating team. None of these changes were substantial (usage criteria as of May 19, 2020 in **eMethods**). The standard tocilizumab dose was 8 mg/kg (maximum 800 mg) x 1; additional doses were discouraged.

During the study period, preference was given to enrollment in the sarilumab RCT. However, given strict trial eligibility criteria and protocol requirements (*e.g*., timed phlebotomy and repeated SARS-CoV-2 testing), tocilizumab was considered in patients ineligible for the trial or when trial enrollment was not feasible due to logistical constraints (*e.g*., outside of enrollment hours or on non-study units). Ultimately, individualized decisions on tocilizumab usage were made by the attending infectious diseases physician and based on many factors including comfort with tocilizumab, concern for bacterial co-infection, and duration on the ventilator.

### Outcomes

The primary outcome was survival probability after intubation. A secondary endpoint assessed status at day 28 on a 6-level ordinal scale of illness severity, including bloodstream infection and pneumonia: (1) discharged alive, (2) hospitalized/off ventilator without superinfection, (3) hospitalized/off ventilator with superinfection, (4) hospitalized/mechanically ventilated without superinfection, (5) hospitalized/mechanically ventilated with superinfection, (6) deceased.

### Covariates

Data were obtained via electronic health record queries and manual abstraction, and included demographics, comorbidities, hospitalization dates, transfer status, laboratory values, microbiology results, concomitant medications, mechanical ventilation dates, oxygenation variables, and discharge status. All positive blood and respiratory cultures were assessed by an Infectious Diseases physician to adjudicate infection versus colonization (TG, LP, SZ, JB). Infections were included if they occurred after intubation and >48 hours after hospitalization. Additionally, only infections occurring after administration of tocilizumab were considered for the treatment arm. For patients who transferred from an outside hospital, length of stay, intubation date, and tocilizumab administration characteristics at that facility were manually abstracted from admission notes. For those intubated at Michigan Medicine, the lowest PaO2/FiO2 ratio in first twelve hours after intubation was also recorded.

### Tocilizumab for COVID-19

Relevant laboratory values at times of presentation and intubation were abstracted. For transfer patients already on mechanical ventilation, the most extreme laboratory values in the first 24 hours after transfer were considered as values at time of intubation. For patients intubated at Michigan Medicine, the most extreme values ± 24 hours from intubation were considered. For patients who received tocilizumab, only laboratory values pre-tocilizumab were considered.

### Other COVID-19 Directed Therapies

Based on available evidence and lack of enrolling clinical trials at local onset of the pandemic, hydroxychloroquine 600 mg every twelve hours x2 doses, then 200 mg every 8 hours was recommended as standard management at the beginning of the study period. Once remdesivir studies were activated, hydroxychloroquine was formally removed from our guidelines on March 26, 2020, and treatment with hydroxychloroquine was rare after these changes. Adjunctive corticosteroid use was generally not recommended, but use in patients with acute respiratory distress syndrome was at the discretion of the critical care physician.

### Statistical Analysis

Descriptive characteristics were provided using means and standard deviations or median and interquartile range for continuous variables, and frequencies and percentages for categorical variables. Kaplan-Meier survival curves were used to describe post-ventilator onset outcomes and time-varying stacked bar plots were applied to demonstrate the 6-level ordinal outcome by elapsed day. Univariate prediction ability of each covariate on the time to death and ordinal outcome at day 28 were explored using Cox proportional hazards models and proportional odds models, respectively. Proportional odds assumption was assessed via Score test. Multiple imputation^19^ was used to impute missing laboratory values for inclusion in sensitivity analyses: twenty-five imputations by fully conditional specification were made based on age, sex, race, ethnicity, transfer status, history of hypertension, congestive heart failure, chronic pulmonary disease, and chronic renal disease. To address non-randomized treatment allocation, we calculated propensity scores by multivariable logistic regression with tocilizumab treatment as the binary outcome and potential confounding factors associated with both outcome and treatment assignment. Using such propensity scores, we applied the inverse probability of treatment weights (IPTW) to create a pseudo study cohort, where the weighted version can balance off the covariate bias and mimic a randomized treatment assignment situation: the IPTW weights for tocilizumab treated patients = 1/p(treated); for untreated patients = 1/[1-p(treated)].^20–22^ All analyses were conducted in univariate and multivariable fashion, and before and after inverse probability of treatment weighting. Sensitivity analyses were performed by thresholds of age, CRP, and D-dimer, and stratified analyses by sex and transfer vs. non-transfers. Analyses were performed in SAS v9.4 and R v4.0.0.

## RESULTS

### Cohort characteristics

Of 484 cases admitted during the study period for COVID-19, 34 were excluded based on enrollment in a sarilumab RCT (NCT04315298). Also excluded were 293 who did not require mechanical ventilation, 2 untreated patients who died within 24 hours of intubation, and 1 infant. Thus, this study included 154 patients requiring mechanical ventilation, including 78 treated with tocilizumab and 76 untreated (**Figure 1**). Median follow-up time was 47 days (range 28–67).

**Figure 1.**
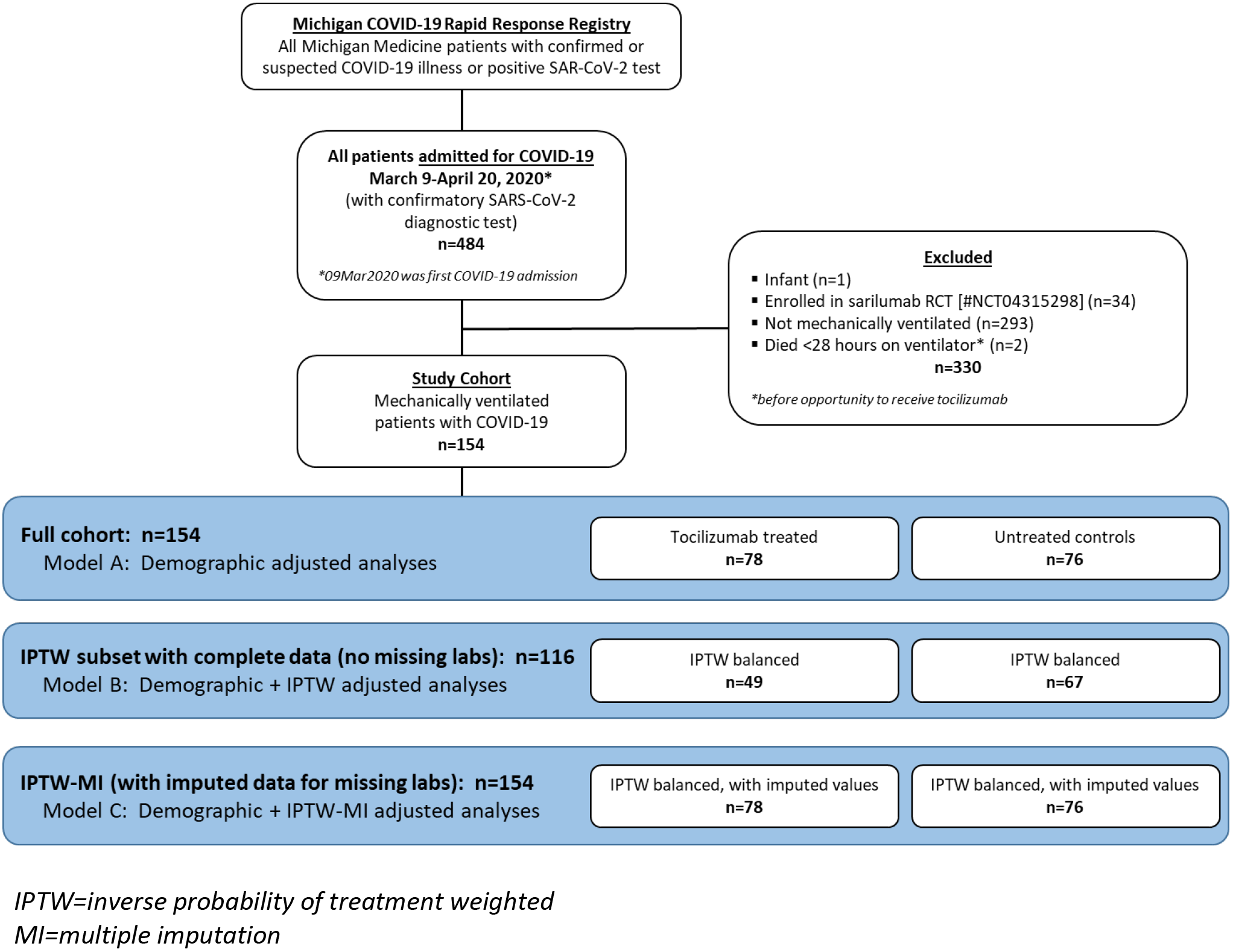
Study cohort flow chart.

Patient characteristics as a function of treatment are shown in **Table 1**. In general, the two groups were well-balanced, and patients were similar with regards to sex, race, most comorbidities, and concomitant therapies. Tocilizumab-treated patients were younger (mean 55 vs. 60 years; p = 0.05) and less likely to have either chronic pulmonary disease (10% vs. 28%; p = 0.006) or chronic kidney disease (35% vs. 49%; p = 0.08). The majority of patients in both groups were transfers from an outside facility, with a higher number of transfers (74% vs. 58%; p = 0.04) in the untreated group.

**Table 1.**
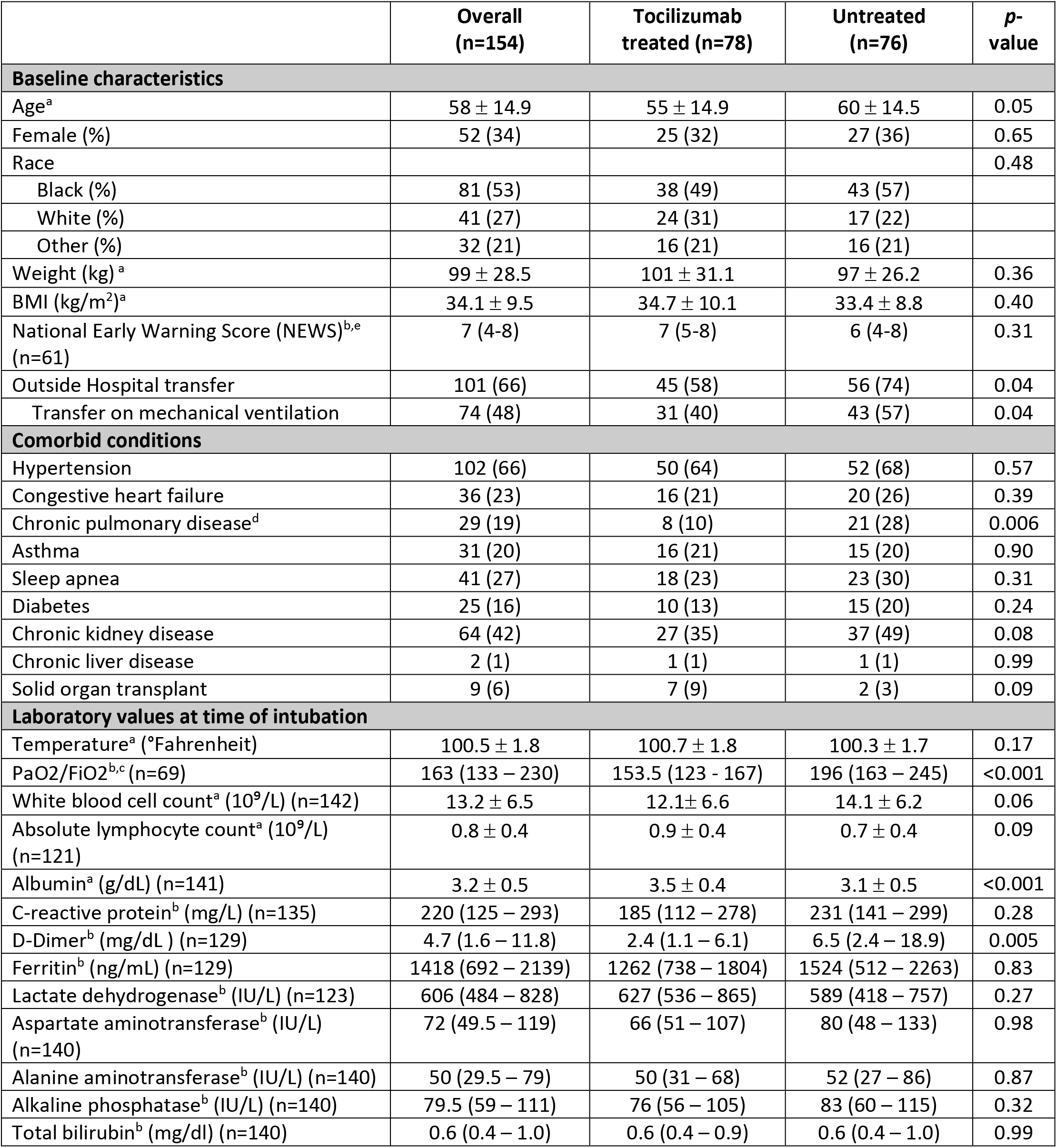
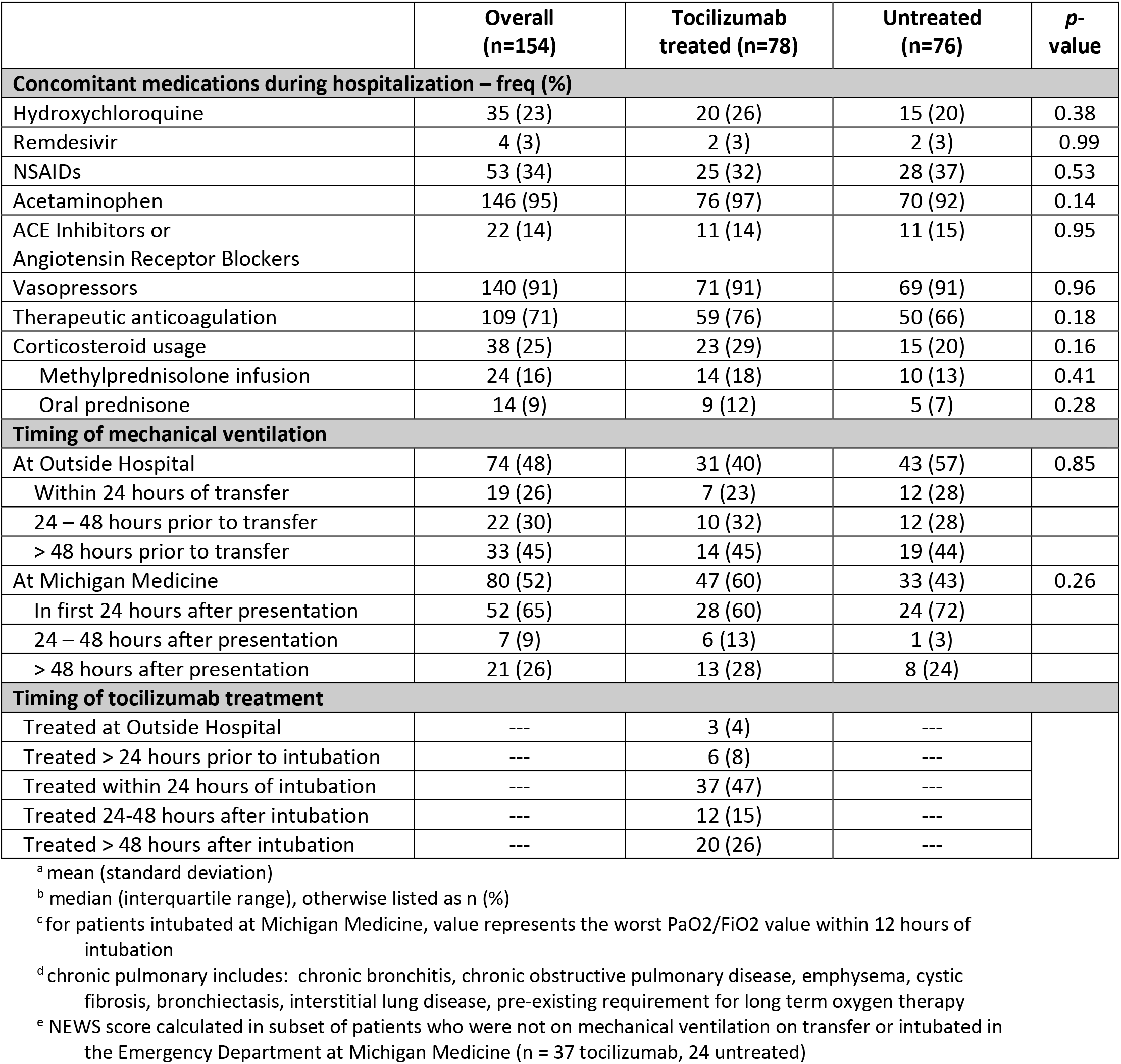
Characteristics of the cohort.

|  | Overall<br>(n=154) | Tocilizumab<br>treated (n=78) | Untreated<br>(n=76) | p-value |
| --- | --- | --- | --- | --- |
| **Baseline characteristics** |  |  |  |  |
| Age<sup>a</sup> | 58 ± 14.9 | 55 ± 14.9 | 60 ± 14.5 | 0.05 |
| Female (%) | 52 (34) | 25 (32) | 27 (36) | 0.65 |
| Race |  |  |  | 0.48 |
| Black (%) | 81 (53) | 38 (49) | 43 (57) |  |
| White (%) | 41 (27) | 24 (31) | 17 (22) |  |
| Other (%) | 32 (21) | 16 (21) | 16 (21) |  |
| Weight (kg)<sup>a</sup> | 99 ± 28.5 | 101 ± 31.1 | 97 ± 26.2 | 0.36 |
| BMI (kg/m<sup>2</sup>)<sup>a</sup> | 34.1 ± 9.5 | 34.7 ± 10.1 | 33.4 ± 8.8 | 0.40 |
| National Early Warning Score (NEWS)<sup>b,e</sup><br>(n=61) | 7 (4-8) | 7 (5-8) | 6 (4-8) | 0.31 |
| Outside Hospital transfer | 101 (66) | 45 (58) | 56 (74) | 0.04 |
| Transfer on mechanical ventilation | 74 (48) | 31 (40) | 43 (57) | 0.04 |
| **Comorbid conditions** |  |  |  |  |
| Hypertension | 102 (66) | 50 (64) | 52 (68) | 0.57 |
| Congestive heart failure | 36 (23) | 16 (21) | 20 (26) | 0.39 |
| Chronic pulmonary disease<sup>d</sup> | 29 (19) | 8 (10) | 21 (28) | 0.006 |
| Asthma | 31 (20) | 16 (21) | 15 (20) | 0.90 |
| Sleep apnea | 41 (27) | 18 (23) | 23 (30) | 0.31 |
| Diabetes | 25 (16) | 10 (13) | 15 (20) | 0.24 |
| Chronic kidney disease | 64 (42) | 27 (35) | 37 (49) | 0.08 |
| Chronic liver disease | 2 (1) | 1 (1) | 1 (1) | 0.99 |
| Solid organ transplant | 9 (6) | 7 (9) | 2 (3) | 0.09 |
| **Laboratory values at time of intubation** |  |  |  |  |
| Temperature<sup>a</sup> (°Fahrenheit) | 100.5 ± 1.8 | 100.7 ± 1.8 | 100.3 ± 1.7 | 0.17 |
| PaO<sub>2</sub>/FiO<sub>2</sub><sup>b,c</sup> (n=69) | 163 (133 – 230) | 153.5 (123 – 167) | 196 (163 – 245) | <0.001 |
| White blood cell count<sup>a</sup> (10<sup>9</sup>/L) (n=142) | 13.2 ± 6.5 | 12.1 ± 6.6 | 14.1 ± 6.2 | 0.06 |
| Absolute lymphocyte count<sup>a</sup> (10<sup>9</sup>/L)<br>(n=121) | 0.8 ± 0.4 | 0.9 ± 0.4 | 0.7 ± 0.4 | 0.09 |
| Albumin<sup>a</sup> (g/dL) (n=141) | 3.2 ± 0.5 | 3.5 ± 0.4 | 3.1 ± 0.5 | <0.001 |
| C-reactive protein<sup>b</sup> (mg/L) (n=135) | 220 (125 – 293) | 185 (112 – 278) | 231 (141 – 299) | 0.28 |
| D-Dimer<sup>b</sup> (mg/dL) (n=129) | 4.7 (1.6 – 11.8) | 2.4 (1.1 – 6.1) | 6.5 (2.4 – 18.9) | 0.005 |
| Ferritin<sup>b</sup> (ng/mL) (n=129) | 1418 (692 – 2139) | 1262 (738 – 1804) | 1524 (512 – 2263) | 0.83 |
| Lactate dehydrogenase<sup>b</sup> (IU/L) (n=123) | 606 (484 – 828) | 627 (536 – 865) | 589 (418 – 757) | 0.27 |
| Aspartate aminotransferase<sup>b</sup> (IU/L)<br>(n=140) | 72 (49.5 – 119) | 66 (51 – 107) | 80 (48 – 133) | 0.98 |
| Alanine aminotransferase<sup>b</sup> (IU/L) (n=140) | 50 (29.5 – 79) | 50 (31 – 68) | 52 (27 – 86) | 0.87 |
| Alkaline phosphatase<sup>b</sup> (IU/L) (n=140) | 79.5 (59 – 111) | 76 (56 – 105) | 83 (60 – 115) | 0.32 |
| Total bilirubin<sup>b</sup> (mg/dl) (n=140) | 0.6 (0.4 – 1.0) | 0.6 (0.4 – 0.9) | 0.6 (0.4 – 1.0) | 0.99 |
| **Concomitant medications during hospitalization – freq (%)** |  |  |  |  |
| Hydroxychloroquine | 35 (23) | 20 (26) | 15 (20) | 0.38 |
| Remdesivir | 4 (3) | 2 (3) | 2 (3) | 0.99 |
| NSAIDs | 53 (34) | 25 (32) | 28 (37) | 0.53 |
| Acetaminophen | 146 (95) | 76 (97) | 70 (92) | 0.14 |
| ACE Inhibitors or<br>Angiotensin Receptor Blockers | 22 (14) | 11 (14) | 11 (15) | 0.95 |
| Vasopressors | 140 (91) | 71 (91) | 69 (91) | 0.96 |
| Therapeutic anticoagulation | 109 (71) | 59 (76) | 50 (66) | 0.18 |
| Corticosteroid usage | 38 (25) | 23 (29) | 15 (20) | 0.16 |
| Methylprednisolone infusion | 24 (16) | 14 (18) | 10 (13) | 0.41 |
| Oral prednisone | 14 (9) | 9 (12) | 5 (7) | 0.28 |
| **Timing of mechanical ventilation** |  |  |  |  |
| At Outside Hospital | 74 (48) | 31 (40) | 43 (57) | 0.85 |
| Within 24 hours of transfer | 19 (26) | 7 (23) | 12 (28) |  |
| 24 – 48 hours prior to transfer | 22 (30) | 10 (32) | 12 (28) |  |
| > 48 hours prior to transfer | 33 (45) | 14 (45) | 19 (44) |  |
| At Michigan Medicine | 80 (52) | 47 (60) | 33 (43) | 0.26 |
| In first 24 hours after presentation | 52 (65) | 28 (60) | 24 (72) |  |
| 24 – 48 hours after presentation | 7 (9) | 6 (13) | 1 (3) |  |
| > 48 hours after presentation | 21 (26) | 13 (28) | 8 (24) |  |
| **Timing of tocilizumab treatment** |  |  |  |  |
| Treated at Outside Hospital | --- | 3 (4) | --- |  |
| Treated > 24 hours prior to intubation | --- | 6 (8) | --- |  |
| Treated within 24 hours of intubation | --- | 37 (47) | --- |  |
| Treated 24-48 hours after intubation | --- | 12 (15) | --- |  |
| Treated > 48 hours after intubation | --- | 20 (26) | --- |  |

<sup>a</sup> mean (standard deviation)
<sup>b</sup> median (interquartile range), otherwise listed as n (%)
<sup>c</sup> for patients intubated at Michigan Medicine, value represents the worst PaO<sub>2</sub>/FiO<sub>2</sub> value within 12 hours of intubation
<sup>d</sup> chronic pulmonary includes: chronic bronchitis, chronic obstructive pulmonary disease, emphysema, cystic fibrosis, bronchiectasis, interstitial lung disease, pre-existing requirement for long term oxygen therapy
<sup>e</sup> NEWS score calculated in subset of patients who were not on mechanical ventilation on transfer or intubated in the Emergency Department at Michigan Medicine (n = 37 tocilizumab, 24 untreated)

Laboratory values at time of intubation are shown in **Table 1**. Tocilizumab-treated patients had lower D-dimer (median 2.4 vs. 6.5 mg/dL; p = 0.005) and higher serum albumin concentrations (mean 3.5 vs. 3.1 g/dL; p< 0.001). Of patients intubated after admission at Michigan Medicine, median PaO2/FiO2 ratios were lower in the tocilizumab group (median 153.5 vs 196; p< 0.001). The timing of mechanical ventilation (**Table 1**) did not differ between the two groups, with the majority of patients being intubated either within 48 hours prior to transfer or during the first 24 hours of admission. Tocilizumab was most commonly administered within 24 hours of intubation, with a minority of use (26%) occurring > 48 hours after intubation.

Propensity score distributions stratified by actual treatment group and diagnostics are shown in **eFigure 1**; odds ratios for tocilizumab receipt by variables included in the propensity score model are in **eTable 3**. Balancing pre- and post-IPTW is shown in **eTable 4**.

### Survival

Survival probability was significantly higher among tocilizumab-treated compared to untreated patients, as displayed by Kaplan-Meier estimates (p = 0.0189); **Figure 2**. Based on Cox proportional hazards models, tocilizumab was associated with a lower hazard of death, after adjusting for demographics [Model A: HR 0.54 (95% CI 0.29, 1.00)], when further IPTW-adjusted for the cohort subset with complete laboratory data [Model B: n = 116, HR 0.55 (0.33, 0.90); IPTW-Kaplan-Meier **eFigure 2**] and when IPTW-MI adjusted (with imputed laboratory data) in the full cohort [Model C: HR 0.54 (0.35, 0.84)]; **Tables 2 and eTable 6**. In stratum-specific sensitivity analyses for transfer patients, direct admits, patients with CRP values > 150 mg/L, D-dimer values > 1.2 mg/dL, and various age cutoffs, similar findings persisted (**eFigure 3, eTable 6**). Case fatality rate at 28 days was also lower for tocilizumab-treated patients (18% vs. 36%; p = 0.01); **Table 2**.

**Table 2.** Outcomes, including superinfections, stratified by treatment.

|  | **Tocilizumab treated**<br>**(n=78)** | **Untreated**<br>**(n=76)** | **p-value** |
| --- | --- | --- | --- |
| 14-day case fatality rate | 7 (9) | 20 (26) | 0.005 |
| 21-day case fatality rate | 11 (14) | 25 (33) | 0.006 |
| 28-day case fatality rate | 14 (18) | 27 (36) | 0.01 |
| Discharged alive by end follow-up | 44 (56) | 30 (40) | 0.04 |
| Length of stay (among discharged)<sup>a</sup> | 20.4 (13.8, 35.8) | 22.9 (16.3, 28.5) | 0.31 |
| Duration of mechanical ventilation<sup>a,b</sup> | 13.8 (7.1, 27.5) | 13.0 (8.1, 23.5) | 0.94 |
| **Hazard ratios for tocilizumab vs control** |  |  |  |
| Model A: demographic adjusted | 0.54 (0.29, 1.00) | ref | 0.05 |
| Model B: demographic + IPTW adj (n=116) | 0.55 (0.33, 0.90) | ref | 0.02 |
| Model C: demographic + IPTW-MI adj | 0.54 (0.35, 0.84) | ref | 0.01 |
| **Odds ratio for proportional odds model for tocilizumab vs control (Day 28)** |  |  |  |
| Model A: demographic adjusted | 0.61 (0.34, 1.09) | ref | 0.10 |
| Model B: demographic + IPTW adj (n=116) | 0.59 (0.36, 0.95) | ref | 0.03 |
| Model C: demographic + IPTW-MI adj | 0.61 (0.40, 0.92) | ref | 0.02 |
| **Superinfection data** |  |  |  |
| Patients with a superinfection | 42 (54) | 20 (26) | <0.001 |
| 28-day case fatality rate<sup>f</sup> | 8 (22) | 5 (28) | 0.61 |
| Patients with pneumonia | 35 (45) | 15 (20) | <0.001 |
| Patients with bloodstream infection | 11 (14) | 7 (9) | 0.34 |
| Time from intubation to first infection<sup>a</sup> | 9.8 (4.5 – 15.8) | 7.7 (3.9 – 14.4) | 0.13 |
| Patients with >1 infection | 10 (13) | 7 (8) | 0.47 |
| **Causative microbiology** |  |  |  |
| **Microbiology of pneumonia<sup>c</sup>** | **n=41** | **n=22** |  |
| *S. aureus* | 21 (51) | 11 (50) |  |
| Methicillin susceptible | 15 (71) | 5 (45) |  |
| Methicillin resistant | 6 (29) | 6 (55) |  |
| *P. aeruginosa* | 5 (12) | 4 (18) |  |
| Multi-drug resistant | 4 (80) | 3 (75) |  |
| *E. coli* | 4 (10) | 1 (5) |  |
| ESBL producing | 1 (25) | 0 |  |
| *K. aerogenes* | 4 (10) | 1 (5) |  |
| *K. pneumoniae* | 3 (7) | 1 (5) |  |
| *S. marcescens* | 3 (7) | 0 (0) |  |
| *S. maltophilia* | 2 (5) | 0 (0) |  |
| Other<sup>d</sup> | 7 (17) | 5 (23) |  |
| **Microbiology of bloodstream infections<sup>c</sup>** | **n=12** | **n=8** |  |
| Coagulase negative staphylococcus | 4 (33) | 3 (38) |  |
| *Enterococcus* spp. | 3 (25) | 2 (25) |  |
| *Candida* spp. | 3 (25) | 1 (13) |  |
| Other<sup>e</sup> | 4 (36) | 2 (28) |  |

<sup>a</sup> Median (interquartile range), otherwise listed as n (%)
<sup>b</sup> limited to those who were extubated alive during the study period (n=94)
<sup>c</sup> There were 41 unique cases of pneumonia in 35 tocilizumab treated patients and 22 unique cases in 15 untreated patients; there were 12 unique bloodstream infections in 11 tocilizumab treated patients and 8 unique bloodstream infections in 7 untreated patients; pathogen numbers can add up to > 100% due to polymicrobial infections
<sup>d</sup> In tocilizumab patients: n=1 *A. baumannii*, *C. koseri*, *C. striatum*, *H. influenzae*, *P. mirabilis*, *P. putida*, and *S. pneumoniae*. In untreated patients: n = 1 *A. fumigatus*, *A. baumannii*, *E. cloacae*, *P. mirabilis*, and *S. pneumoniae*.
<sup>e</sup> In tocilizumab patients n=1 Methicillin susceptible *S. aureus*, *S. mitis*, *E. coli*, and *K. pneumoniae*; in untreated patients n = 1 Methicillin resistant *S. aureus* and *E. cloacae*.
<sup>f</sup> Limited to patients with infection in first 28 days: 37 tocilizumab treated vs. 18 tocilizumab untreated

**Figure 2.**
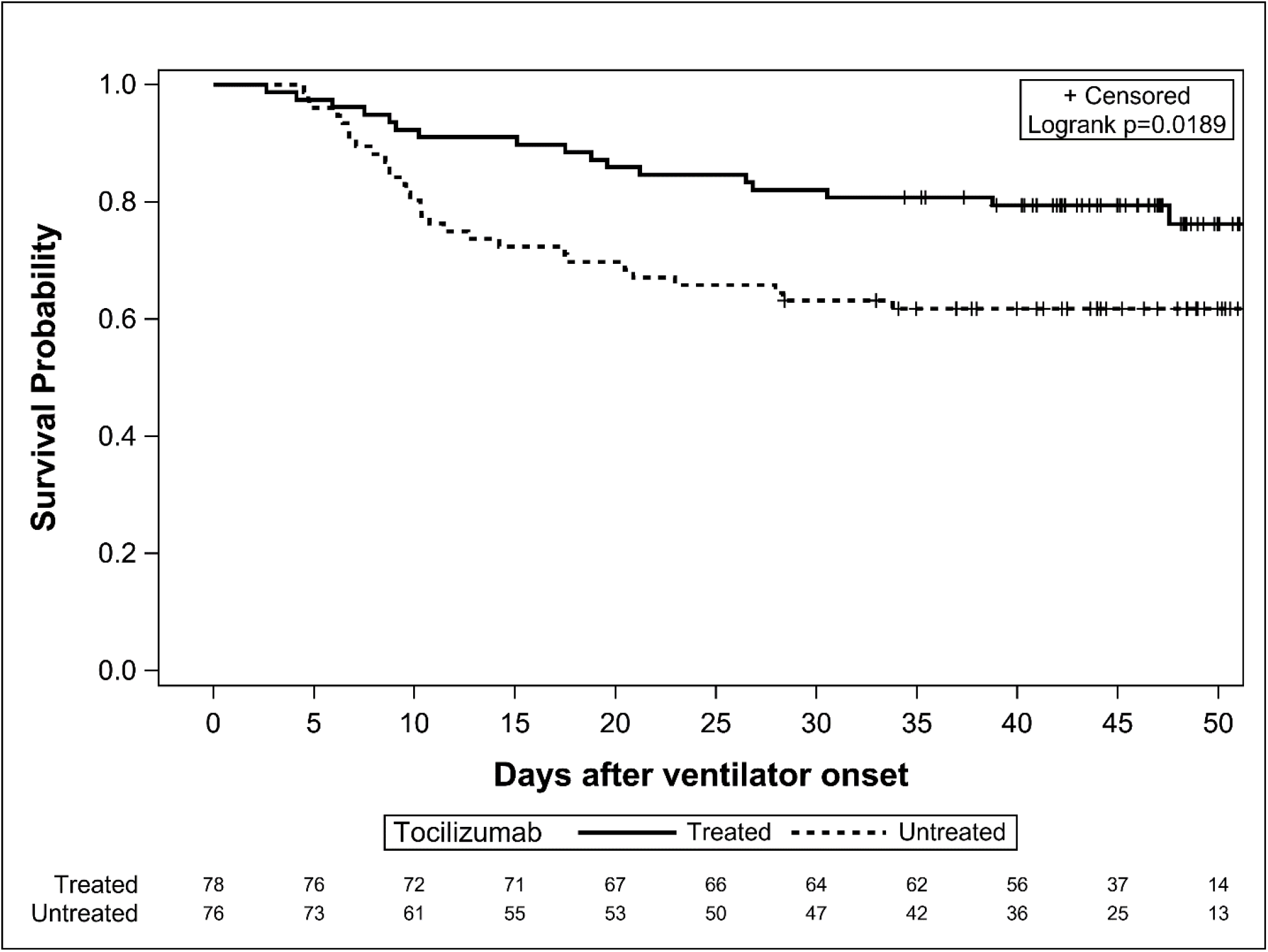
Kaplan-Meier estimates for probability of survival as a function of time since mechanical ventilation onset, stratified by tocilizumab treatment (n = 154; n = 46 deaths).

Figure 3A & 3B.

Patient status post-ventilator onset on a six-level ordinal scale integrating superinfection occurrence, stratified by tocilizumab treatment.

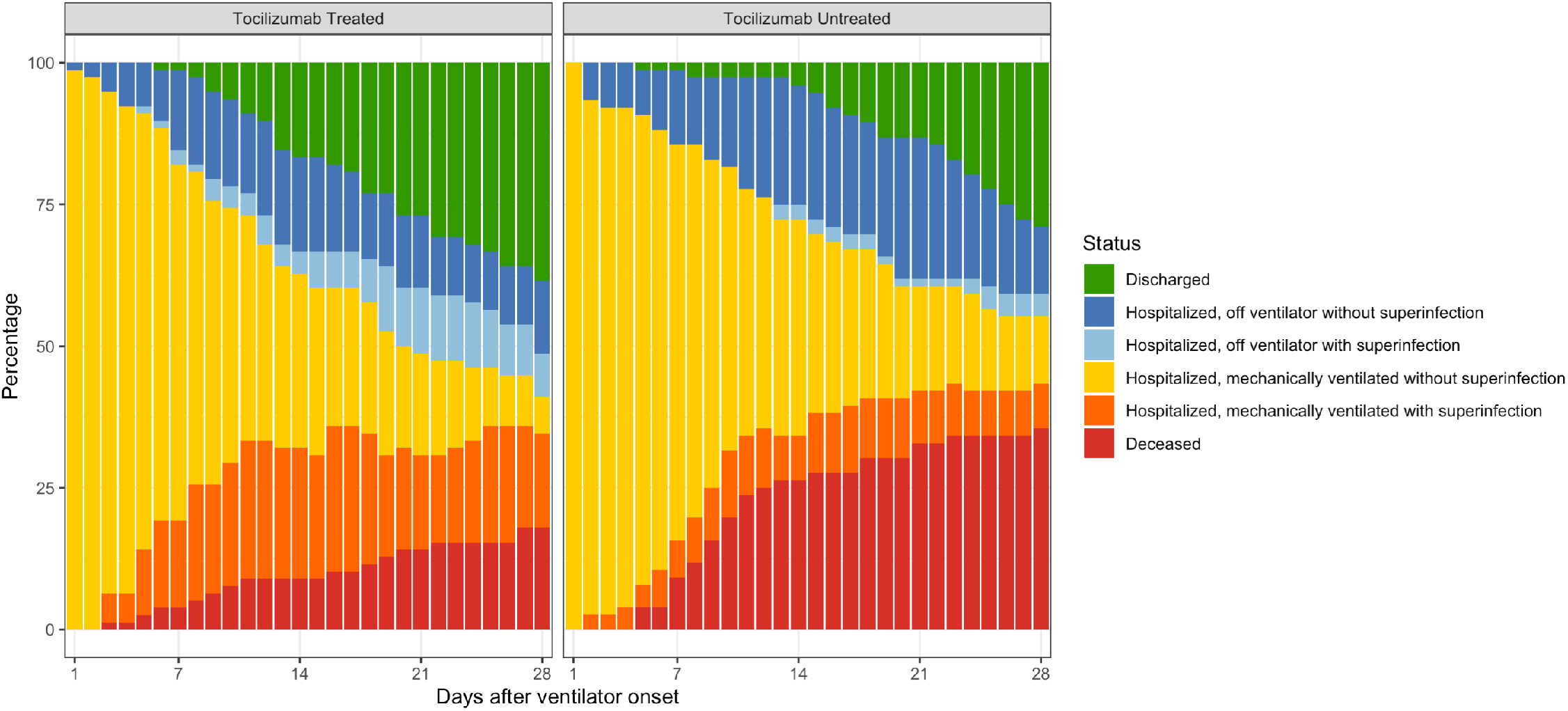
The distribution of patient status, by number of days after onset of mechanical ventilation through day 28 of follow-up.

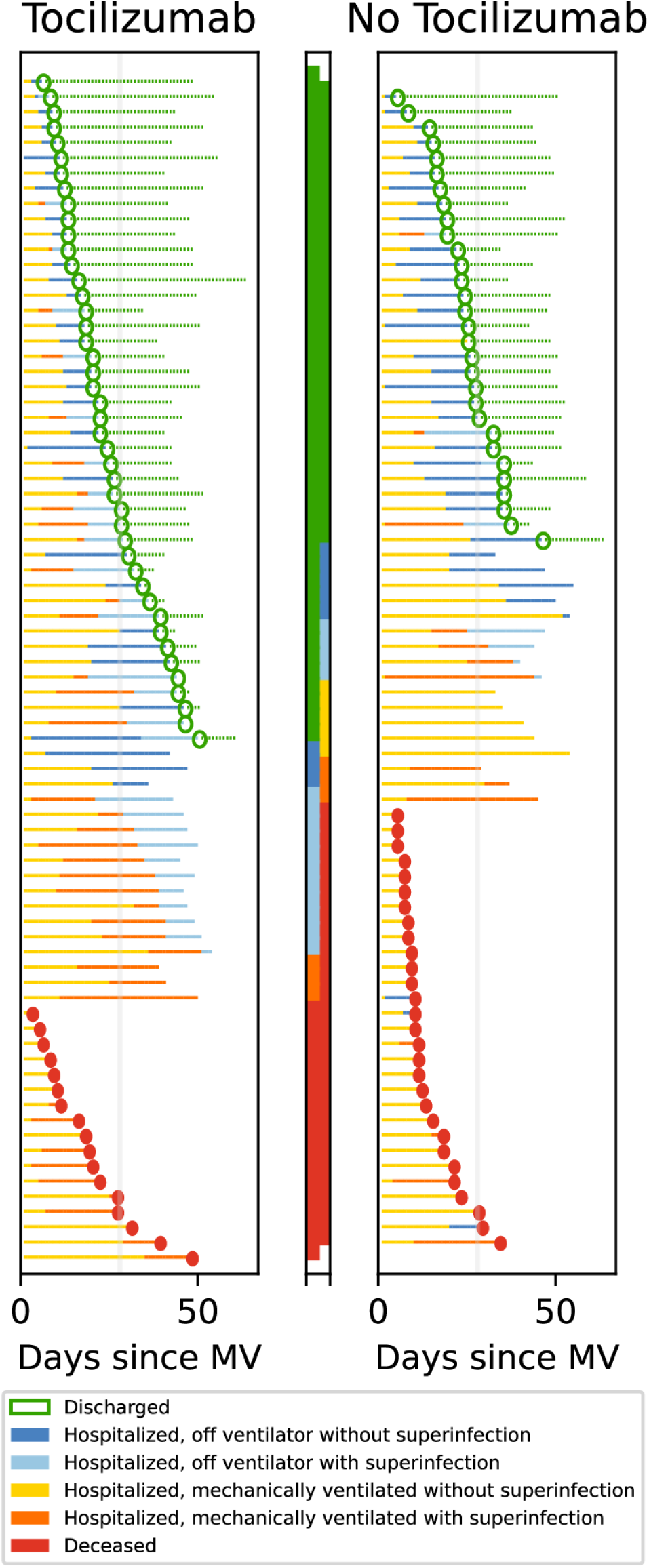
Individual patient trajectories on the six-level ordinal scale over the study period. Each row represents changes in individual patient status from time of onset of mechanical ventilation until event (death) or end of the study period (May 19, 2020). Horizontal lines correspond to elapsed time, with colors corresponding to clinical status of the patient. Solid circles represent death, and hollow circles represent discharge from hospital (alive). The middle panel indicates the most recent patient status. Grey vertical lines mark 28-day follow-up.

### Superinfections

Patients who received tocilizumab were more than twice as likely to develop a superinfection than untreated controls (54% vs. 26%; p< 0.001), driven primarily by a large increase in ventilator-associated pneumonia (45% vs. 20%; p< 0.001); **Table 2**. There was no difference between groups with regards to timing of infection, incidence of bloodstream infections, or development of more than one infection. The causative microbiology of superinfections was similar between groups. *Staphylococcus aureus* accounted for ∼50% of the bacterial pneumonias in both arms. Case fatality rates at day 28 were similar among tocilizumab-treated patients who had a superinfection and those who did not [8/37 (22%) vs. 6/41 (15%); p = 0.42].

### Ordinal Outcome Integrating Effectiveness and Infection Data

Stratified by treatment arm, **Figure 3A** depicts the daily distribution of status on the six-level ordinal scale through day 28, while **Figure 3B** displays individual patient trajectories. Tocilizumab administration was associated with improved status in the demographic- and IPTW-adjusted proportional odds models (OR per 1 level increase in outcome scale): Model A/demographic-adjusted: 0.61 (0.34, 1.09); Model B/demographic + IPTW: OR 0.59 (0.36, 0.95); Model C/demographic+IPTW-MI: OR 0.61 (0.40, 0.92)] (**Tables 2 & eTable 7, eFigure 4**). Furthermore, patients who received tocilizumab were more likely to be discharged alive over the study period (56% vs. 40%; p = 0.04).

## DISCUSSION

In this observational, controlled study of patients with severe COVID-19 necessitating mechanical ventilation, receipt of tocilizumab was independently associated with improved survival. Importantly however, tocilizumab was also associated with increased incidence of secondary bacterial pneumonia and bloodstream infections. While this did not appear to negatively influence ultimate outcome, and case fatality rates were similar in infected and uninfected tocilizumab-treated patients, this finding highlights the need for adequately powered randomized controlled trials further evaluating efficacy and safety of tocilizumab in COVID-19.

Respiratory failure in severe COVID-19 is frequently characterized by high serum IL-6 concentrations.^23^ Excessive IL-6 can induce lung epithelial cells to increase inflammatory responses, leading to increased macrophage response and ultimately pulmonary damage. IL-6 may also be a significant contributor to thrombosis, having been associated with both tissue and vascular endothelial cell injury, and contributing to platelet aggregation and angiotensin II microvascular dysfunction.^24,25^ Conversely, as a critical cytokine in organizing T-cell responses to infections, IL-6 may play a beneficial role in COVID-19. It may suppress viral reactivation,^26^ protect against superinfection, and facilitate lung repair and remodeling *after* viral injury.^27^ Thus, our approach was to administer tocilizumab in patients who were rapidly desaturating or recently intubated, in an attempt to optimize the timing of administration for maximal benefit. Our dosing strategy (single, high dose of 8 mg/kg) was an attempt to saturate receptors to rapidly inhibit IL-6 signaling but also allow more rapid clearance in order to not interfere with tissue remodeling and limit immunosuppression.

Our results support these hypotheses. Given the heterogeneity in tocilizumab treatment decisions between providers at our institution, the two groups in this analysis were largely comparable with regard to factors impacting COVID-19 outcomes. Although there were slight imbalances with regards to age, baseline D-dimer, CRP, and transfer status, we utilized rigorous methods for observational data accounting for these factors and treatment propensity. Tocilizumab remained associated with better outcomes across modeling strategies. Furthermore, results remained consistent in sensitivity analyses limiting to patients with D-dimer > 1.2 mg/dL (HR 0.42) or CRP > 150 mg/L (HR 0.48) values above thresholds previously associated with mortality (preprint);^5^ **eTable 6, eFigure 3**. Likewise, the protective effect associated with tocilizumab persisted when restricted to various age groups (< 60, < 70 or < 75 years, HRs 0.55–0.59), patients transferred from an outside facility (HR 0.54), or direct admits (HR 0.41).

In addition to the survival advantage, receipt of tocilizumab was associated with a significant improvement on a six-point ordinal scale that incorporated mechanical ventilation, development of superinfection, and discharge from the hospital (OR 0.61, 95% CI 0.40, 0.92). This improvement in illness severity level with receipt of tocilizumab is reflected in the statistically significant increase in patients discharged home over the study period (56% vs. 40%; p = 0.04), and the finding that of the 17 patients remaining in the hospital at the end of follow-up, only 3 (18%) of tocilizumab-treated patients remained on the ventilator, compared to 8/17 (47%) of untreated controls (**Figure 3B**). This consistent advantage across the ordinal scale strengthens the confidence in the benefit demonstrated with tocilizumab in this cohort and furthermore has significant resource conservation implications.

Importantly, these data also reinforce concerns with superinfection risk due to IL-6 inhibition. To date, the risk of superinfection in mechanically ventilated patients with severe COVID-19 remains poorly described and the incremental risk associated with a single dose of tocilizumab is not well characterized. We demonstrated that superinfection was common in this population, with 39% developing a pneumonia or bloodstream infection. Furthermore, tocilizumab was associated with higher occurrence of infection (54% vs. 26%; p< 0.001), driven primarily by the development of ventilator-associated bacterial pneumonia in 45% of patients receiving tocilizumab. Interestingly, we also identified an association between severe COVID-19 infection and staphylococcal pneumonia, as roughly half of the cases in both the tocilizumab and control group were due to *S. aureus*.

Although these data are observational, several strengths of the study warrant comment. First, this analysis utilizing a Rapid Response Registry informed by an internationally-designed clinical characterization protocol,^16^ represents the first well-controlled, comparative analysis assessing safety and effectiveness of tocilizumab for severe COVID-19. In order to address potential confounding by indication or other imbalances between groups, propensity scoring and multivariable models were utilized, as well as sensitivity analyses. Across various analytic strategies, results consistently indicated benefit associated with tocilizumab. Additionally, median follow-up time for the cohort was 47 days (range 28–67), with all patients followed for at least 28 days, representing a substantially longer observation period than many COVID-19 treatment studies to date, and indicative of sustained benefit. Furthermore, all secondary infections were reviewed by an infectious diseases physician to ensure accurate reporting.

However, this analysis is not without limitation. First and foremost, randomized controlled trial data will be critical for confirming the perceived benefits from this observational study and better quantify risks. Second, there were incomplete data for laboratory variables, though we used contemporary methods for imputing missing data. Additionally, we focused on the impact of tocilizumab 8 mg/kg x 1 in mechanically ventilated patients. This study does not address the potential role of tocilizumab earlier in illness for preventing mechanical ventilation, the optimal dose of tocilizumab, the potential utility of multiple doses, or the role of IL-6 serum concentrations (which were not routinely available) in predicting tocilizumab response, all of which are important questions that warrant further investigation.

In conclusion, tocilizumab was associated with improved survival, despite higher occurrence of superinfections, in a cohort of COVID-19 patients requiring mechanical ventilation. These data are encouraging and can help to inform clinical practice while results from randomized controlled trials of IL-6 inhibitors are awaited.

## Data Availability

Data contain PHI and only accessible to IRB-approved investigators

## ACKNOWLEDGMENTS

This work was supported by the National Institutes of Health (NIH/NCATS UL1TR002240; NIH/NHLBI 1K12HL133304 to CMF); the Centers for Disease Control and Prevention (CDC U01IP000974); and an ASTCT New Investigator Award (to JLG). The findings and conclusions in this report are those of the authors and do not necessarily represent the official position of the NIH, CDC or the Department of Health and Human Services.

